# Medical Findings in Infants Prenatally Identified with Sex Chromosome Trisomy in Year One of Life

**DOI:** 10.1101/2024.07.10.24310206

**Authors:** Nicole Tartaglia, Shanlee Davis, Susan Howell, Samantha Bothwell, Kayla Nocon, Karen Kowal, Chijioke Ikomi, Andrew Keene, Victoria Reynolds, Agnethe Berglund, Judith Ross

## Abstract

**BACKGROUND AND OBJECTIVE:** Sex chromosome trisomies (SCT), including XXY, XYY, and XXX syndromes, have been historically underdiagnosed. Noninvasive prenatal cell-free DNA screening has significantly increased identification of these conditions, leading to a need for pediatric care for a growing population of newborns with SCT. Our goal was to analyze and compare perinatal features, medical diagnoses, and physical features in infants with prenatal identification of SCT conditions through the first year of life.

**METHODS:** The eXtraordinarY Babies Study is an ongoing, prospective natural history study of prenatally identified children with SCT conducted by interdisciplinary teams in Colorado and Delaware. Participants were enrolled prior to 12 months of age and had pregnancy, birth, medical histories, and physical exams completed by board-certified pediatricians at 2, 6, and/or 12-month visits. Descriptive statistics were followed by comparisons between SCT groups using t-tests or ANOVA, Fisher exact, and correlations between medical features with alpha of 0.05. Relative risks were calculated compared to general population rates.

**RESULTS:** 327 infants were included in the analysis (XXY=195, XXX=79, XYY=53). Major congenital anomalies were rare (1.7%). Relative risk compared to general population was elevated for breastfeeding difficulties (51.7%;RR 2.7), positional torticollis (28.2%;RR 7.2), eczema (48.0%;RR 3.5), food allergies (19.3%;RR 2.4), constipation requiring intervention (33.9%;RR 7.6), small cardiac septal defects (7.7%;RR 17.2), and structural renal abnormalities (4.4%;RR 9.7). Inpatient hospitalization was required for 12.4%, with 59.5% of hospitalizations attributable to respiratory infections.

**DISCUSSION:** These findings of medical conditions with a higher prevalence can inform anticipatory guidance and medical management for pediatricians caring for infants with SCT.

**Article Summary:** Medical findings in largest cohort of prenatally identified infants with XXY, Trisomy X, and XYY from birth to 12 months and implications for pediatric care.

*What’s Known on This Subject:* One in ∼500 individuals have an extra X or Y chromosome, or sex chromosome trisomy (SCT). Prenatal screening is now routinely identifying SCT, however there are few studies to guide perinatal and infant care for these individuals.

*What This Study Adds:* This prospective observational study presents medical features for 327 infants with prenatally identified SCT from birth through the first year of life. Results identify where proactive screenings and/or interventions may be warranted for infants with SCT.

## INTRODUCTION

Sex chromosome trisomies (SCT) occur in approximately 1:500 births and include XXY (Klinefelter syndrome) and XYY syndrome in males, and XXX (Trisomy X) syndrome in females. ^1,2^ Historically, prenatal diagnosis of SCT occurred only when chorionic villi sampling (CVS) or amniocentesis was indicated, such as due to abnormal ultrasound findings or advanced maternal age, accounting for only 5-10% of all cases of SCT. ^3^ An additional 5-25% were diagnosed postnatally upon presentation of clinical symptoms, however 65-90% of SCT cases remained undiagnosed. ^3^ The adoption of cell-free DNA (cfDNA) screening into routine prenatal care in the US has led to a marked increase in the prenatal identification of SCT, ^4^ resulting in a large cohort of infants with a known diagnosis. With a median of over 4,000 visits annually per pediatric physician, ^5^ each pediatrician will have several patients with SCT in their practice.

Due to the previously low rate of diagnosis in the prenatal period, there is a paucity of research on medical features in the early years of life. Prior international newborn screening studies identified 200 infants with SCT in the 1960’s. These reports suggest that most infants with SCT are born at term and are not at an increased risk of congenital malformations or medical conditions during the neonatal period. ^6,7^ In childhood, tall stature and mild hypotonia are commonly described in the literature, as well as reports of atopic conditions, autoimmunity, constipation, seizure disorders, tremor, and recurrent respiratory infections. ^7–12^ However, the preexisting literature relies on small sample sizes and selection bias, and therefore may not be generalizable to the population identified through prenatal cfDNA screening. Furthermore, while the medical complexity of these children is often assumed to be low, there is no information on healthcare utilization, such as hospitalization or surgery. Prevalence data for medical comorbidities collected prospectively are needed to guide best care practices for pediatric SCT care and provide appropriate counseling to parents.

The surge of infants with SCT being identified by prenatal cfDNA screening provides a rich opportunity for prospective study of these infants to inform the natural history of these conditions. The eXtraordinarY Babies Study collects comprehensive data on health and development for prenatally identified infants with SCT starting within the first year of life to better inform least-biased risk and guide counseling and care for these infants. The aim of this study is to describe the prospective physical and medical features in the perinatal period and first year of life for a large, prenatally ascertained cohort of infants with SCT, and to compare features between SCT conditions and to the general population estimates.

## METHODS

Parents of infants identified prenatally and subsequently confirmed to have SCT were invited to participate in an IRB-approved natural history study called the eXtraordinarY Babies Study at one of two sites in Colorado or Delaware (ClinicalTrials.gov NCT03396562; COMIRB 17-0118; Nemours IRB# 1151006). Comprehensive descriptions of study methodology including recruitment and assessment measures have been previously published and are summarized in Figure 1, and this report focuses on medical and physical features identified within the first year of life. ^13^ In brief, inclusion criteria include genetic testing by amniocentesis or postnatal cytogenetic testing confirming SCT with less than 20% mosaicism with a typical cell line, gestational age at or above 34 weeks at birth, and enrollment prior to 12 months of age. Following written informed consent, data were collected by medical providers using review of medical records and a standardized interview with the parent(s) developed for the study that included detailed birth and medical histories. Study visits occurred at 2, 6, and/or 12-months of age depending on timing of enrollment, and visits were conducted in-person and/or via telehealth due to COVID-19 restrictions during the study period.

**FIGURE 1:**
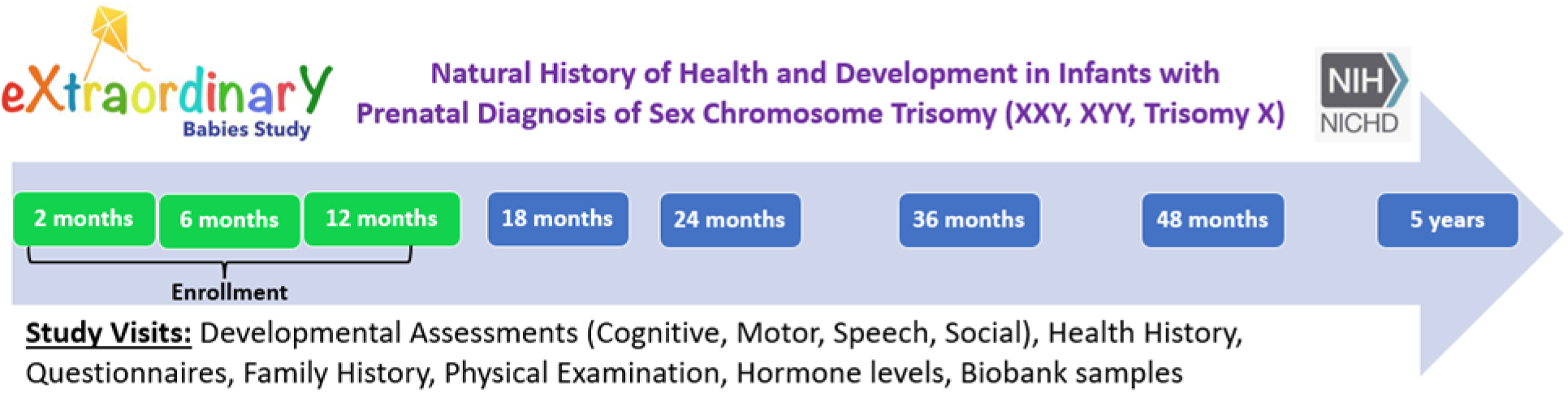
Schematic of eXtraordinarY Babies Study Visits.

Demographic data were collected through a standardized parent survey, and 4 Factor Hollingshead Index was calculated as previously described. ^14^ Physical examination was conducted by board-certified pediatrician at 2, 6, and/or 12-month visits in person visits. Birthweight (BW) and length (BL) measurements were converted to z-scores for gestational age and sex according to US norms. ^15^ Descriptive metrics (proportions, means) and estimates (95% confidence intervals or standard deviations) for each outcome were calculated for the whole sample as well as stratified by SCT. Outcomes were compared between SCT groups using fisher’s exact tests, for low cell counts, or ANOVA followed by pairwise comparisons. Finally, when rigorous published data were available for the general pediatric population, risk ratios and 95% confidence intervals were calculated by unconditional maximum likelihood estimation (Wald) in the epitools R package to determine whether the SCT sample proportion differed from what is expected in the general population. All analyses were two-sided with a type 1 error rate set at 0.05. Data were stored and managed in the secure REDCap database and all analyses were conducted in RStudio v2022.12.0 using R v.4.2.2.

## RESULTS

The full sample consisted of 327 infants (XXY n=195, XXX n=79, XYY n=53) representing 47 US states (Figure 2) and diverse racial and ethnic backgrounds (Table 1). Nearly all (95%) were initially identified to have SCT via prenatal cfDNA screening performed either for advanced maternal age or no clinical indication (elective), while the minority (∼5%) had abnormal ultrasound and/or biochemical markers prompting prenatal genetic testing (Table 2). All participants except for 3 females with XXX were non-mosaic; the 3 mosaic cases were 47,XXX/46,XX with <20% 46,XX cell line in all cases. Pregnancies were largely uncomplicated, and the vast majority of the cohort was born full-term via vaginal delivery at a mean gestational age 38.7±1.5 weeks. Most infants (85.2%) were average-for-gestational age (AGA), however, infants with XXY and XXX had lower mean weight and length z-scores at birth than those with XYY.

**FIGURE 2:**
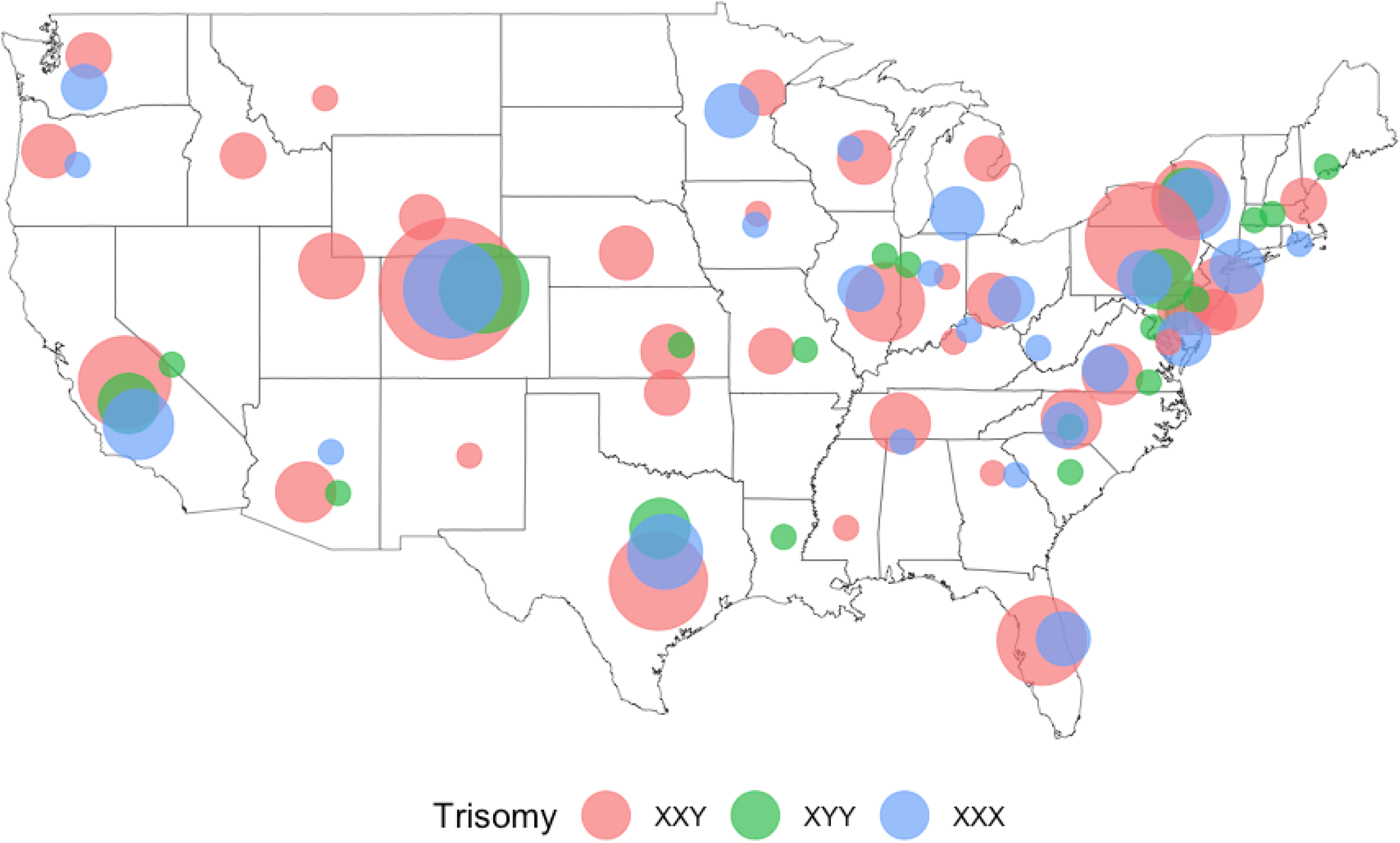
Participant US Regional Representation. Point Sizes range from N = 1 to N = 38. Hawaii and Alaska are both represented with N = 1. N = 9 patients are international.

**TABLE 1:**
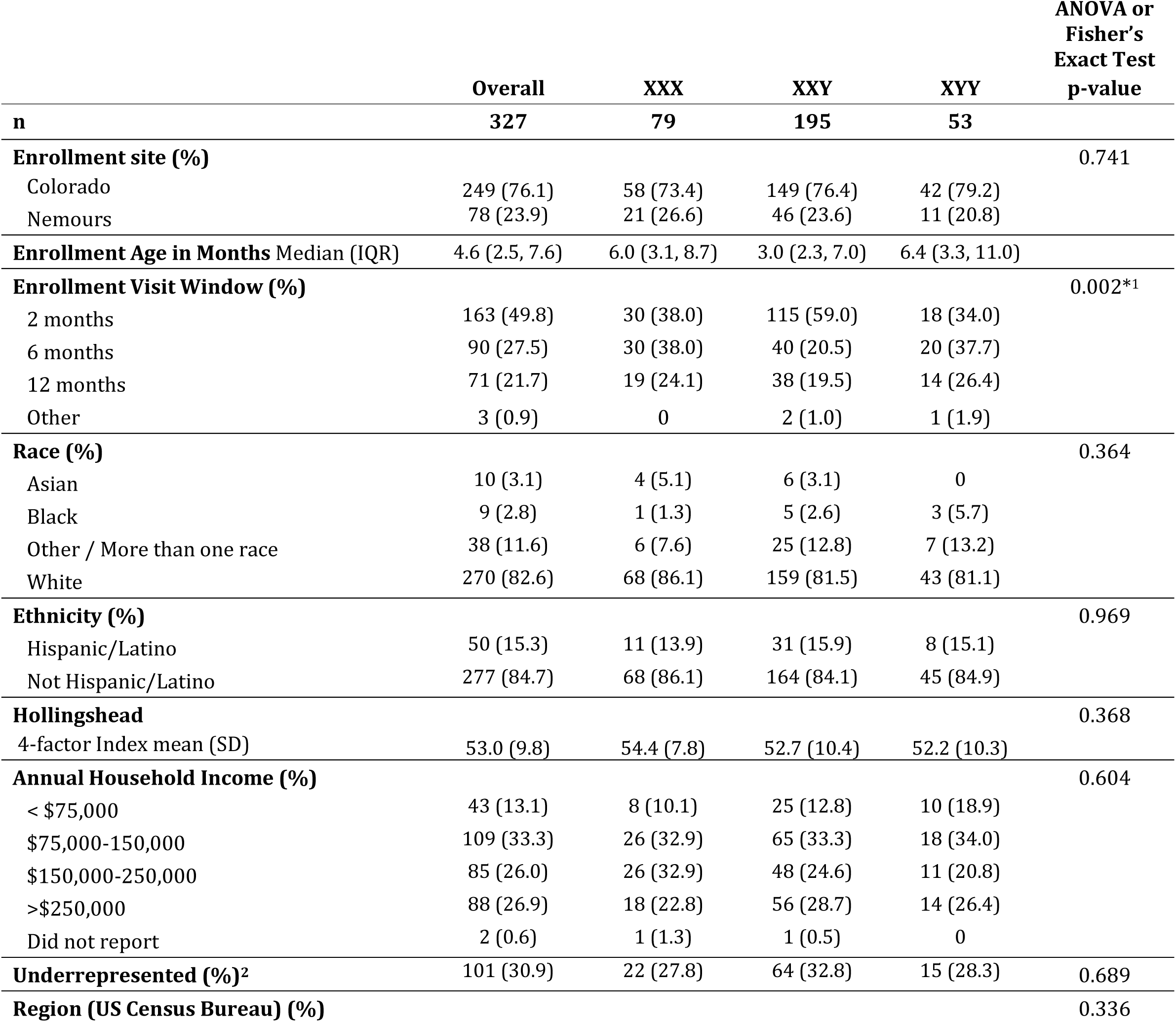
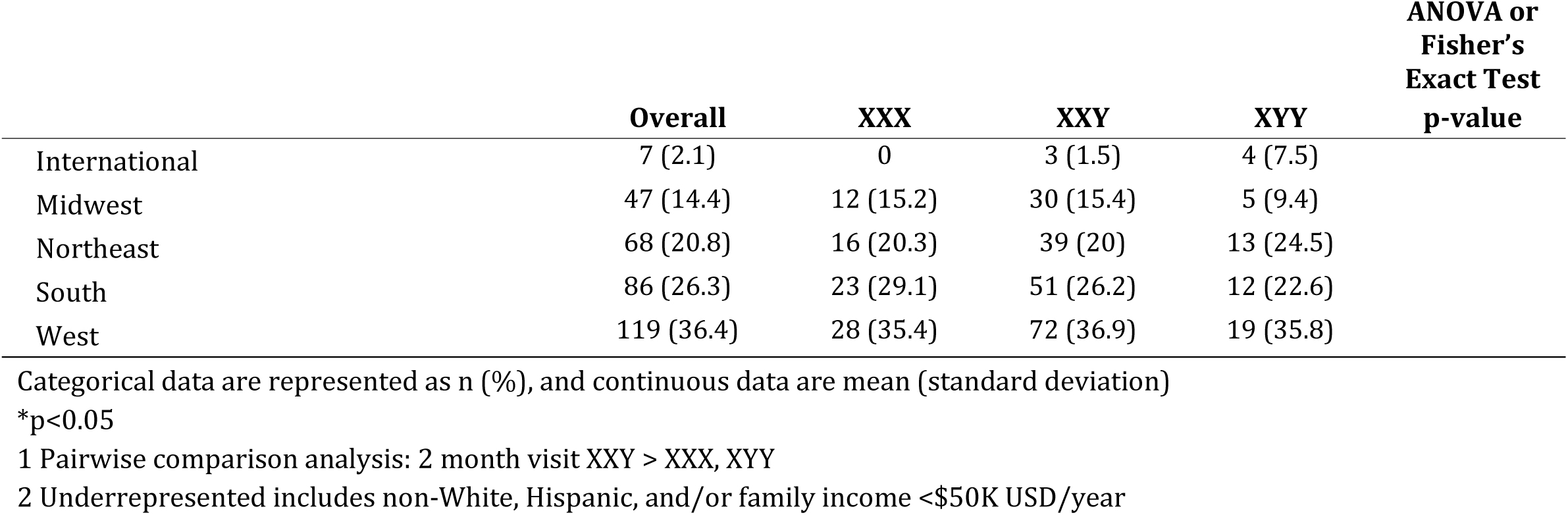
Participant Demographics.

**TABLE 2:**
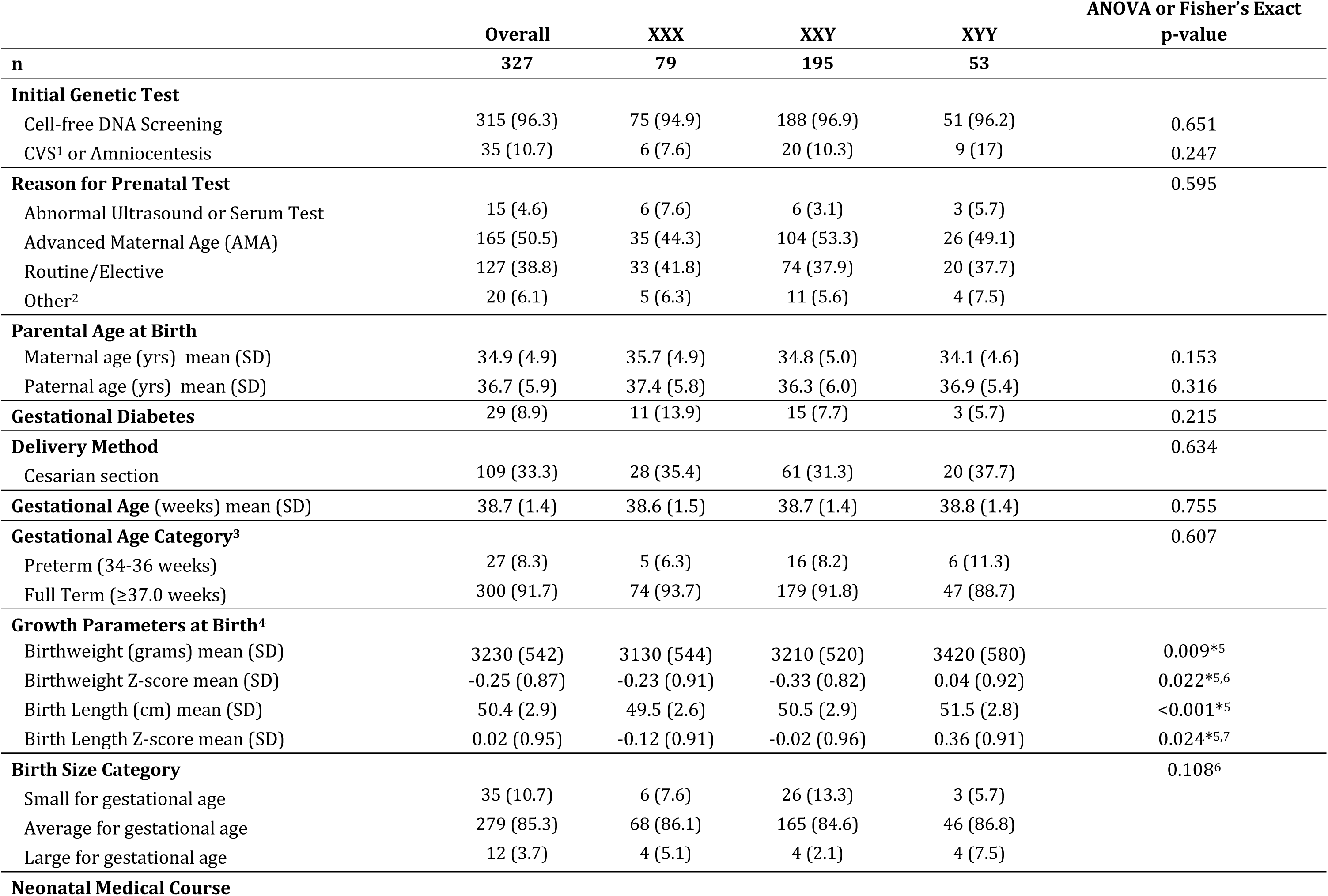
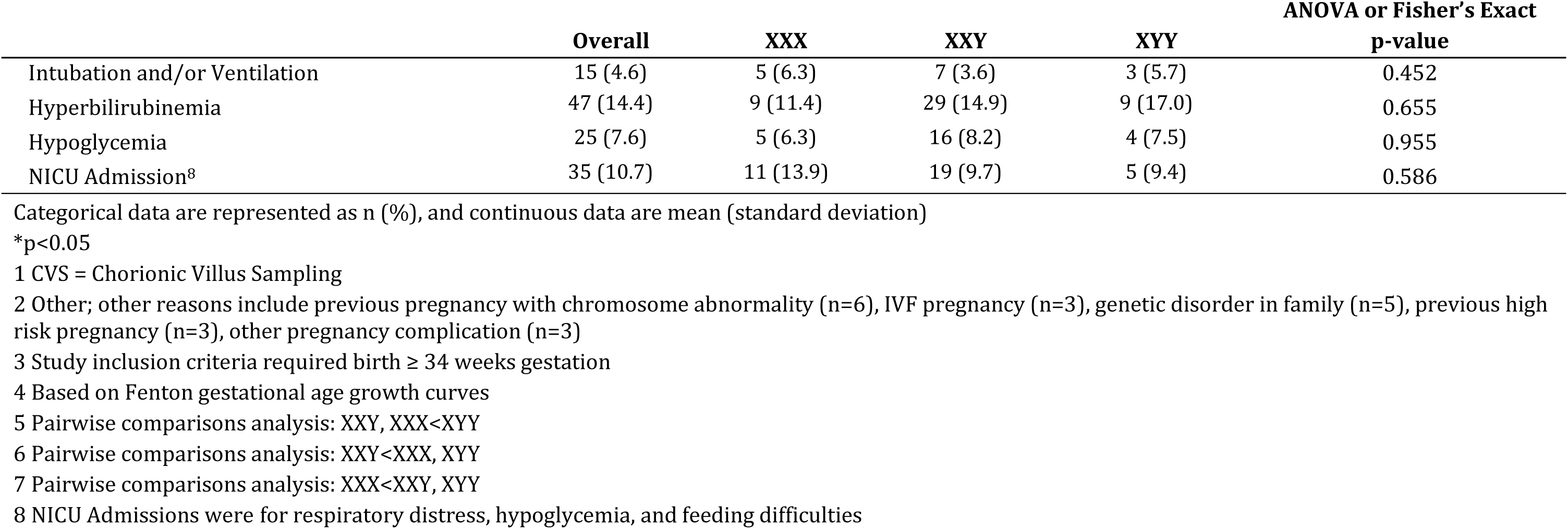
Prenatal and Perinatal History.

Major congenital anomalies were identified in 1.7% of the cohort, including hypospadias (n=3, all subcoronal), cleft lip/palate (n=1), and clubfoot (n=1), which is not different from general population estimates (up to 3%^16^) (Table 3). Common minor congenital anomalies (Figure 3) were fifth digit clinodactyly (49.0%), epicanthal folds (45.1%), torticollis (28.2%), plagiocephaly (18.8%), umbilical hernia (9.1%), and preauricular tags (6.3%). Overall mean z-score for intercanthal distance was nearly a full standard deviation higher than norms (z-score –0.94 (95% CI: -1.37, 2.79), however only 2.7% met criteria for hypertelorism (z-score > 2.0). There were very few differences in the rates of physical findings between SCT groups. Males with XXY had lower stretched penile length compared to XYY at 2 months (p<0.001), however mean length in the XXY group was only a quarter of a standard deviation below the mean for the general population, and only 8 (5.4%) met criteria for short penis length (stretch penile length z-score <-2.0) in those with a study exam who had not received testosterone treatment (Table 4).

**FIGURE 3:**
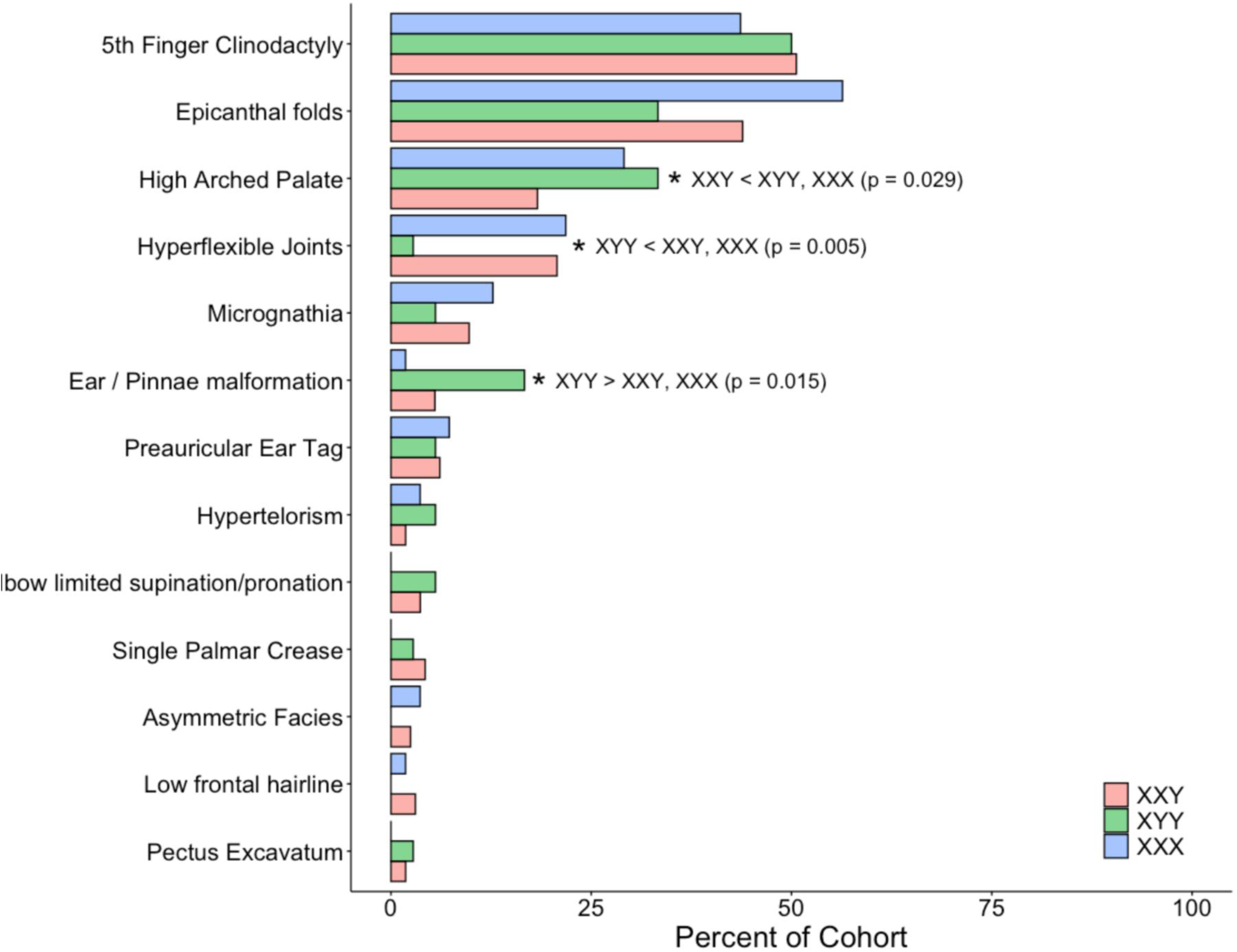
Physical Features on Examination.

**TABLE 3:**
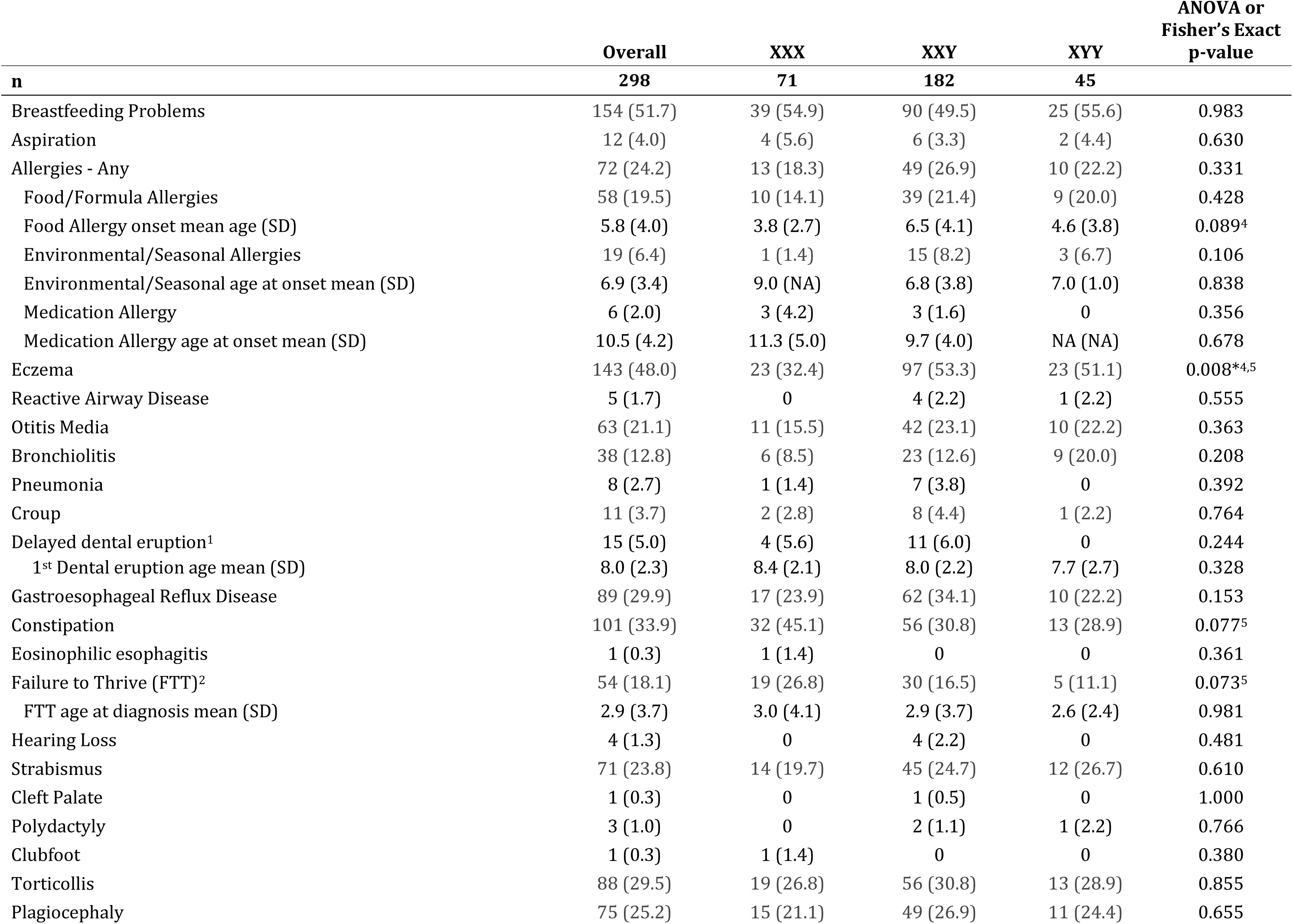
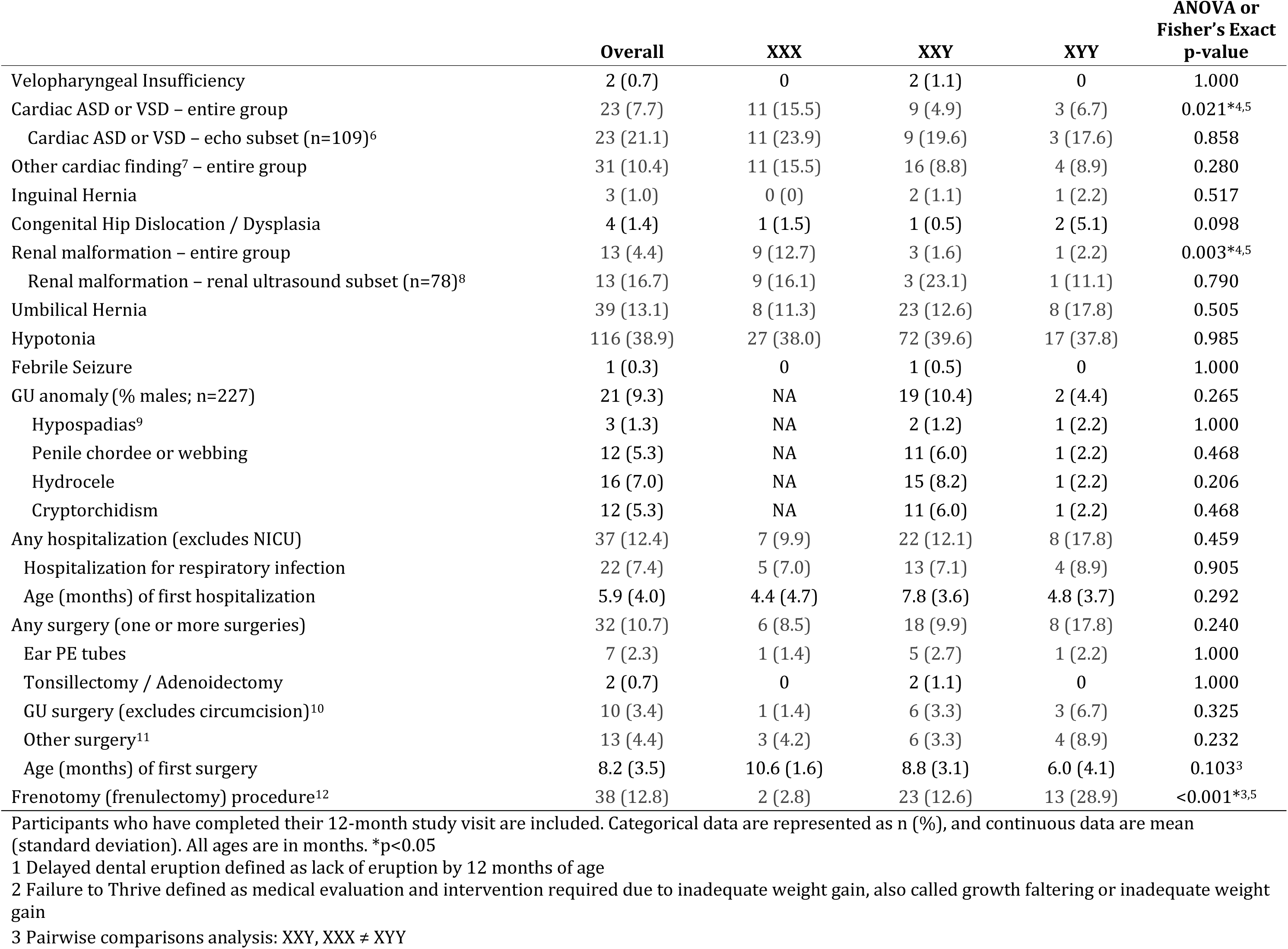
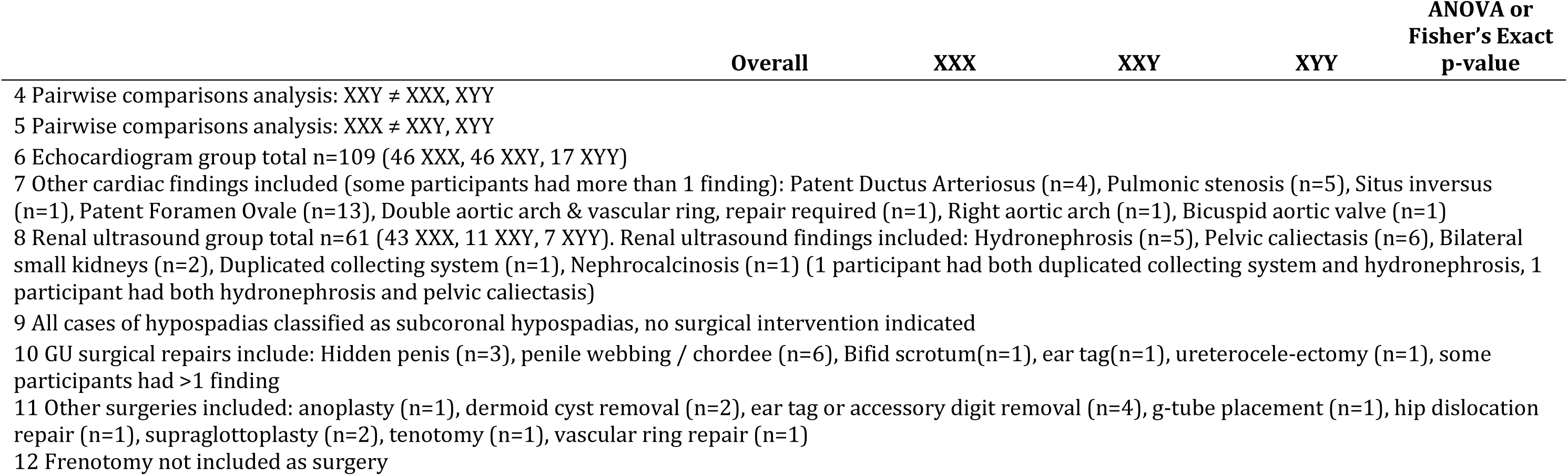
Medical Problems from Birth to 12 months.

**Table 4.**
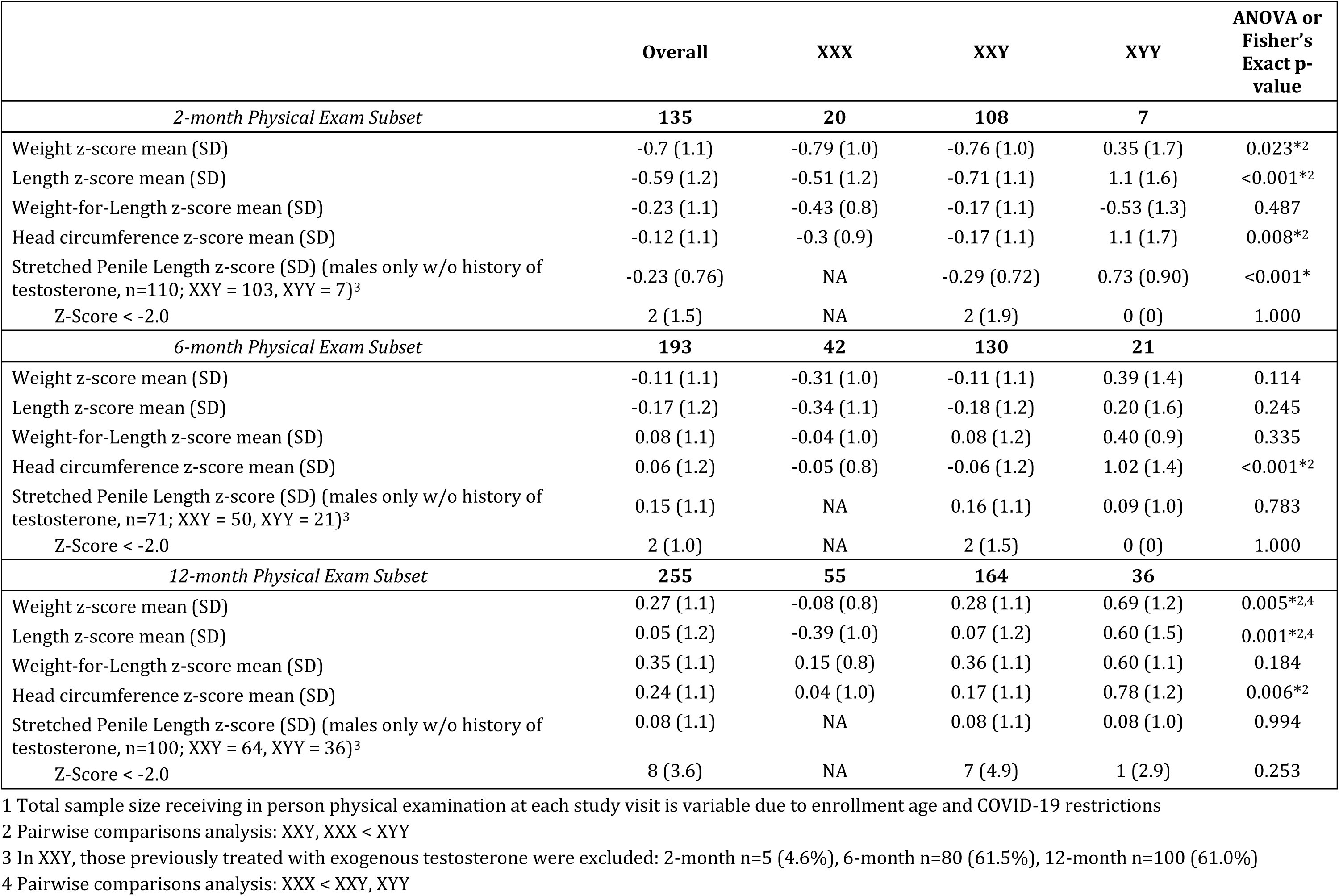
Physical Examination Growth z-scores at 2, 6, and 12 months^1^.

Study protocol did not include standard imaging, however of those who had clinical postnatal echocardiograms completed (n=109), cardiac septal defects (atrial and/or ventricular) were found in 23 (21.1%), which is 7.7% of the cohort overall. None of the infants with identified ASD or VSD required invasive cardiac procedures within the first year of life, however one infant required surgery for a symptomatic vascular ring secondary to double aortic arch. Of the 78 infants who had a clinical renal ultrasound, 13 (16.7%) had a structural abnormality, which represents 4.4% of the full cohort (vs 0.45% in the general population, ^17^ p<0.001). When stratified by karyotype, girls with XXX were more likely to have received an echocardiogram and/or renal ultrasound, however the proportion with abnormal findings of those who had imaging did not significantly differ between SCT subtypes.

Breastfeeding difficulties occurred in half of the cohort and equally between the three SCT conditions, predominantly due to difficulties with latch and/or inadequate milk transfer. Despite this, the mean duration of breastmilk feeding was 8.0±5.5 months with 61.1% of infants still receiving breastmilk at 6 months and 43.3% at 12 months, both at or above US national statistics (58.2% and 37.6%, respectively). ^18^ The presence of breastfeeding difficulties was associated with receiving a frenotomy (21.7% vs 4.8%, p<0.001), earlier age of formula introduction (3.6±3.5 vs 4.7±3.5 months, p=0.013), as well as a diagnosis of failure to thrive (28.0% vs 11.9%, p=0.002). There was no relationship with breastfeeding difficulties and hypotonia, micrognathia, high arched palate, reflux, or constipation (p>0.05 for all). Constipation was more common in all SCT groups compared to the general population (33.9% vs 7.0%, p<0.001) ^19^ and was associated with hypotonia (p<0.001).

Atopic conditions were present in 54.7% of all infants within the first year of life. Eczema was present in 48.0% of all infants with SCT vs 13.7% in general population (RR 3.5 [3.1-3.9], p<0.001) ^20^ and allergies (food, environmental, and/or medication) were present in 24.2% vs 10.4% in the general population (RR 2.6 [2.1-3.2], p<0.001) ^21^. The risk of food allergies was 2.4 times greater in infants with SCT compared to general population estimates (19.3% vs 8%, 95% CI 1.9-3.1, p<0.001) ^22^, with a mean age of 5.8±4.0 months at diagnosis. Food allergies were more common among infants with eczema (28.7% vs 11.0%, p=0.001) and reflux (31.6% vs 13.7%, p<0.001).

Acute infectious diagnoses including otitis media (21.1%), bronchiolitis (12.8%) and pneumonia (2.7%) were similar to estimates in the general population. Overall, 12.4% of the cohort required inpatient hospitalization within the first year of life, with 59.5% of these hospitalizations attributable to respiratory etiologies. Surgeries occurred in 10.7% of the cohort, with the most common indication being for repair of minor GU anomalies.

Weight and weight-for-length z-scores increase in the first year of life for all SCTs, while length z-scores increase from birth to the first year of age for boys but not XXX (Table 4 and Figure S1). By 12 months of age, growth parameters in all 3 groups were within 1 standard deviation of the mean for the general population.

## DISCUSSION

This large, prospective cohort study describes the comprehensive natural history of prenatal, birth, physical findings and medical conditions arising within the first year of life for infants prenatally identified to have SCT. Overall, major congenital anomalies and complex medical conditions were uncommon and there were minimal differences in perinatal and infant findings between SCT groups. We observed several findings warranting clinical consideration within the first year of life, including breastfeeding problems, failure to thrive, torticollis, constipation, structural cardiac and renal differences and atopic conditions (Table 5). The results from this study directly inform pediatric care as providers and families can be reassured that a prenatal diagnosis of SCT is not associated with complex medical or physical abnormalities within the first year of life, but proactive monitoring for select at-risk conditions is warranted.

**TABLE 5:**
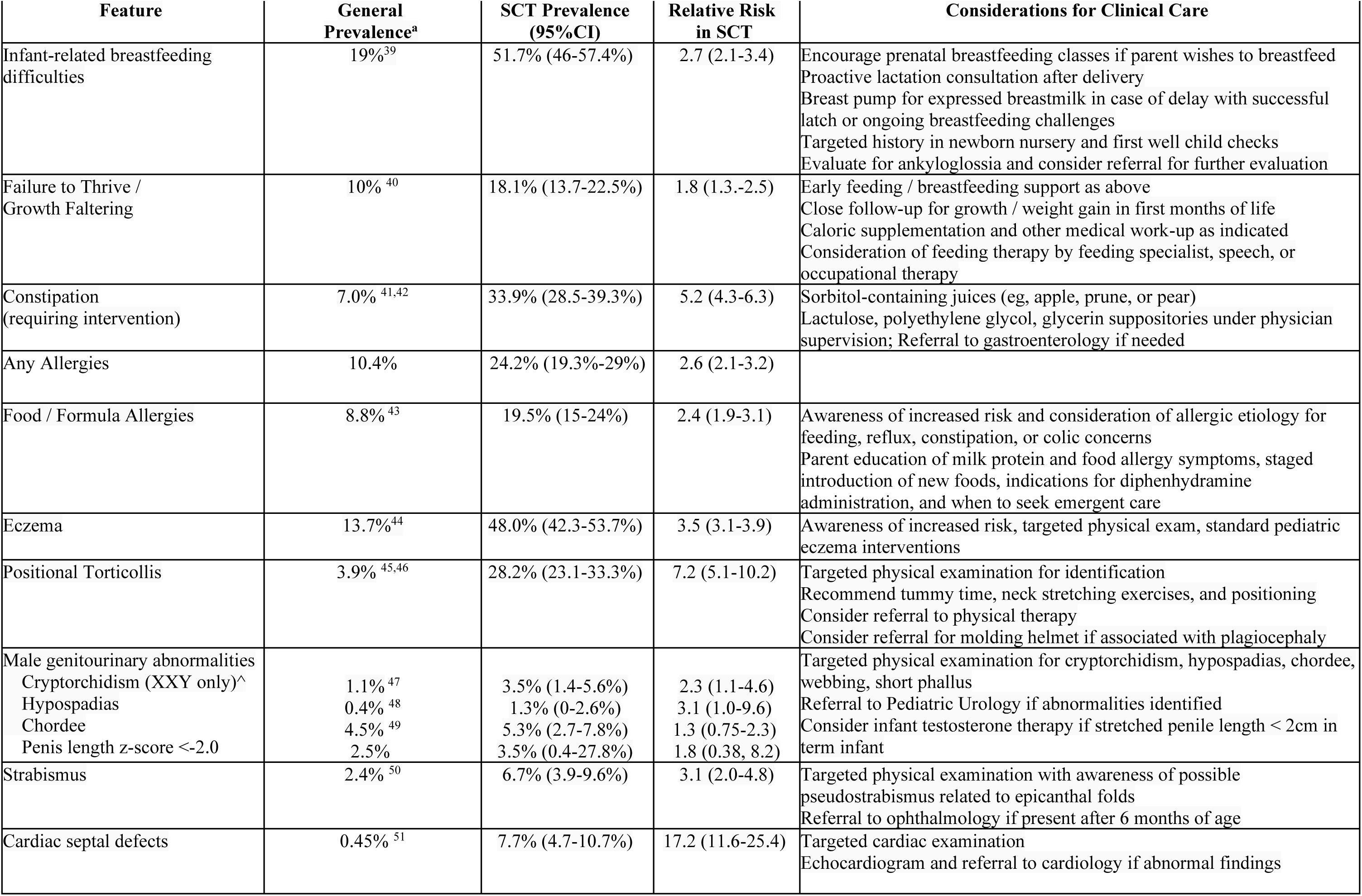
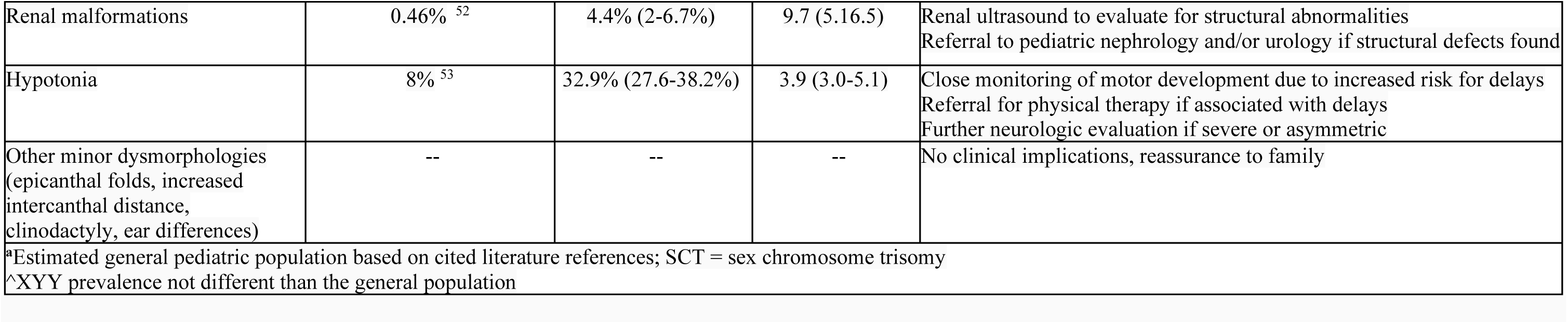
Medical Conditions at Higher Risk in Infants with Prenatal SCT Diagnosis and Guidance for Clinical Care.

Our results support that pregnancies affected by SCT are generally healthy and neither fetal nor maternal complications are obviously different from the general population. Based on these observations, standard prenatal care is appropriate, aside from genetic counseling and decisions on timing of diagnostic genetic testing. ^23^ However, as conditions requiring medical interventions occurred in approximately 1 out of 4 neonates in our cohort, we recommend that delivery is planned at a facility equipped to manage potential neonatal complications including hypoglycemia, hyperbilirubinemia requiring phototherapy, and respiratory insufficiency. Furthermore, our observed prevalence of transient neonatal hypoglycemia may be an underestimates as most term, AGA infants are not routinely evaluated for hypoglycemia and neonatal hypoglycemia often does not present with overt symptoms. ^24^ Therefore, neonates with SCT may benefit from routine glucose screening and additional research.

While major congenital malformations were rare, structural cardiac and renal anomalies occurred more commonly than expected and this prevalence is likely underestimated given the minority of participants had echocardiograms or renal imaging. Indications for echocardiogram or renal imaging varied, with some being performed as general screening due to a known chromosomal abnormality and others due to prenatal concern or postnatal exam findings such as murmur. Congenital cardiac and renal anomalies have been previously associated with XXX, ^25^ but these conditions in males with SCT are primarily limited to case reports. ^26–30^ The high prevalence of abnormal imaging among all infants with SCTs supports universal postnatal screening with echocardiogram and renal ultrasound to identify anatomical defects, however it is also important to acknowledge that none of the individuals required intervention within the first year of life unless symptoms were present. Longitudinal follow up and additional studies are needed to confirm whether structural cardiac and renal imaging findings in SCTs have clinical implications warranting universal imaging in asymptomatic individuals.

Interestingly, in males with XXY, structural cardiac and renal defects were just as common as genital abnormalities which traditionally have been more commonly associated with XXY. While cumulatively ∼10% of the XXY cohort did have minor genitourinary differences which have previously been attributed to testosterone differences in XXY, penile structural abnormalities (hypospadias and chordee) were also observed in 4.4% of males with XYY, suggesting a possible etiology associated with the additional sex chromosome rather than a hormonal etiology. Regarding penile size, true micropenis on our examination was rare, with mean penile length z-score in XXY of -0.3 in those not treated with testosterone. This is important as there is common belief that XXY is associated with short penile length. While a thorough GU examination is advised, most babies will have typical genitalia. Subsequent analyses will evaluate serum hormone concentrations and relationship to health, physical, and developmental outcomes.

Despite a highly motivated study sample with >90% attempting breastfeeding, breastfeeding difficulties were present in over half of the sample and associated with inadequate growth for a subset in the first months of life. We did not systemically evaluate quality of breastfeeding, reasons for breastfeeding challenges, or ankyloglossia which warrant further study. We noted a high rate of frenotomy procedures with differences between karyotypes (29% in XYY). Ankyloglossia occurs in ∼7% of infants in the general population^31^ and is a known cause of breastfeeding difficulties. Further, frenotomy can be beneficial if ankyloglossia is present. ^32–33^ The high rate of frenotomy observed in our cohort highlights the need for additional research to identify the true prevalence and classifications of ankyloglossia in infants with SCT, the actual contribution to feeding problems, and whether frenotomy directly improves breastfeeding. Breastfeeding challenges may reflect neurodevelopmental delays in oral-motor planning and coordination, congruent with previous studies that have identified oral-motor dyspraxia in SCTs. ^34–37^ Other anatomical differences (such as micrognathia or high-arched palate), ^38^ low muscle tone, or poor endurance could also theoretically contribute to feeding difficulties, however we did not observe a relationship with these features and feeding problems in our cohort. The relationship of early feeding problems with later developmental outcomes will be explored in future analyses as the cohort ages. From a clinical perspective, proactive support with lactation consultation may be beneficial, and it is important to appreciate that many infants with SCT were able to receive breastmilk for a similar duration as the general population, either through adequate lactation support and/or expressed breastmilk. Studies of the etiology of breastfeeding problems and specific interventions to support successful breastfeeding in this population are needed.

The prevalence of atopic conditions was one of the most striking medical findings occurring in the first year, with nearly half having eczema and 1 in 5 with a food allergy. This association has been previously reported in SCTs, although we did not expect to find this high of prevalence within the first year of life. While we did not systematically capture disease severity, anecdotally eczema was rarely severe and improved with age. Nevertheless, these findings suggest an altered immune system response in infants with SCT that warrants additional investigation.

We acknowledge limitations of the study, including sample size discrepancies between SCT groups, subjectivity of some of the outcomes, and reliance on published reference data for comparison to the general population. Overestimation of some findings may occur given a diagnosis of a genetic condition may introduce bias to search for and potentially identify abnormalities. It is also possible that our study underestimates the true prevalence of congenital anomalies in SCTs as our study does not include fetuses that may have been spontaneously or electively terminated. Recognizing these limitations, this study contributes vital statistics describing early phenotypic features in SCTs and has important counseling and clinical management implications.

As this cohort ages through the toddler and school-aged years, comprehensive prospective data collection will allow for broader descriptions on the natural history of medical features into childhood. Ongoing recruitment priorities include increased racial, ethnic, and socioeconomic diversity of the study cohort and more balanced sample sizes between the three SCT conditions. Analyses exploring the relationships of family medical history, neurodevelopmental outcomes, reproductive hormone concentrations, and other systemic biomarkers may identify specific predictors or subpopulations most at risk for poorer health outcomes that would guide more intensive early supports, therapies, and medical follow-up.

As prenatal cfDNA screening is now routinely identifying SCT conditions and more infants are presenting to primary care providers with a prenatal diagnosis, there is an acute need for contemporary evidence on perinatal and infant comorbidities to guide counseling and care. Our results support that counselors and medical providers can reassure families that most infants have a typical perinatal course, and that most medical conditions that arise are common in the general pediatric population, well-known to pediatric providers, and respond to evidence-based interventions. For those medical conditions that are indeed more common and cause challenges in the first year of life, anticipatory guidance and close monitoring can facilitate appropriate interventions and timely support.

## Data Availability

All data produced in the present study will be available through NICHD DASH

## Conflict of Interest Disclosures (includes financial disclosures)

The authors have no conflicts of interest to disclose.

## Clinical Trial Registration and Data Sharing Statement (if any)

This study is registered on ClinicalTrials.gov NCT03396562 https://clinicaltrials.gov/study/NCT03396562. Data sharing is available through requests from NICHD DASH (https://dash.nichd.nih.gov/) and includes deidentified individual participant data, study protocols, DASH data codebook, and the informed consent form. The data will be made available to researchers who provide methodologically sound proposals approved by the NICHD DASH Data or Biospecimen Access Committee.

## Abbreviations

AGA: Average for gestational age
AMA: Advanced maternal age
ASD: Atrial septal defect
BW: Birth weight
BL: Birth length
CDC: Centers for Disease Control
COMIRB: Colorado Multiple Institutional Review Board
CVS: Chorionic villus sampling
GU: Genitourinary
LGA: Large for gestational age
NICU: Neonatal Intensive Care Unit
PE: Pressure equalizing
SCT: Sex chromosome trisomies
SGA: Small for gestational age
VSD: Ventricular septal defect

## Contributors Statement Page

Drs. Nicole Tartaglia and Shanlee Davis conceptualized and designed the study, designed data collection instruments, supervised data collection, collected data, developed data analysis plan, drafted the initial manuscript, and critically reviewed and revised the manuscript.

Dr. Judith Ross conceptualized and designed the study, supervised data collection, collected data, and critically reviewed and revised the manuscript.

Dr. Chijioke Ikomi, Dr. Agnethe Berglund, Susan Howell and Karen Kowal collected data and critically reviewed and revised the manuscript.

Samantha Bothwell conducted data analysis, generated tables and figures, and critically reviewed and revised the manuscript.

Kayla Nocon and Victoria Reynolds participated in participant recruitment, study visit coordination, data collection and management, and critically reviewed and revised the manuscript.

Andrew Keene participated in data collection, data sharing, manuscript preparation, and critically reviewed and revised the manuscript.

All authors approved the final manuscript as submitted and agree to be accountable for all aspects of the work.

## ACKNOWLEDGEMENTS

We would like to acknowledge additional team members contributing to this study including previous research coordinators Amira Herstik, Tanea Tanda, Sierra Kaiser, Caroline Harrison, Jillian Kirk, Mariah Brown, Amanda Alstrom, Gabrielle Stefy, and Julia Jaen. We would like to thank AXYS (www.genetic.org), Living with XXY, and the Colorado Genetic Counseling Symposium, for support in recruitment and study dissemination, the ACMG Newborn Screening Translational Research Network team for support with database design and data sharing.

---

**FIGURE S1:**
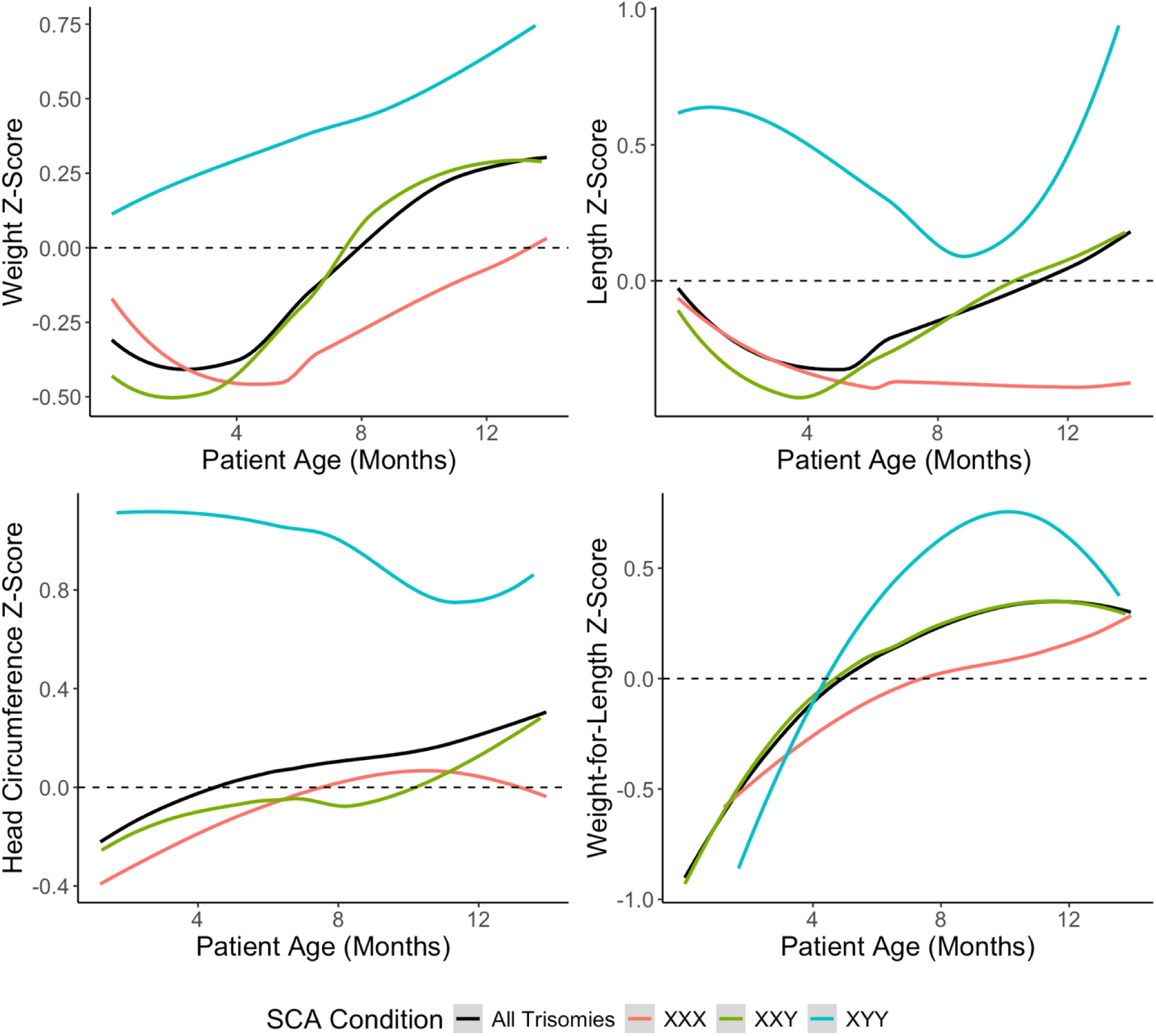
Participant Growth Z-Score Trajectories. LOESS (locally estimated scatterplot smoothing) smoothed trajectories of growth parameter z-scores by trisomy condition

